# Patient friendly summaries of oncology consultations generated by large language models - A pilot study of patient and provider satisfaction

**DOI:** 10.1101/2025.10.13.25337951

**Authors:** Sonali Harchandani, Ryann Quinn, Kriti MIttal, Alex Martin, Ming-Jin Wang Ryan Holstead

**Author notes:** Corresponding Author: Ryan Holstead.

## Abstract

The expanding capacity of large language models allow for improvements in patient and provider healthcare quality and experience. The medical oncology consultation often includes a discussion of a life-limiting diagnosis and complex treatment protocols. Patient recall from the discussion may be limited, and it is possible that a patient specific written summary could help with understanding, recall, and overall experience. Using a privacy compliant large language model, a prompt was instructed to rewrite an ambulatory medical consultation note as a patient friendly summary, capturing key details from a diagnosis and treatment plan. The summary was provided to both provider and patient for review and a 5-point Likert survey was administered inquiring on the output’s accuracy, clarity, and helpfulness. Patients reported agreement in 100%, 100%, and 87% on each topic respectively. 93% of patients recommended the use of similar summaries in the future. Providers reported agreement in 98%, 91%, and 96% for accuracy, clarity, and empathy respectively. All providers (100%) recommended similar summaries to be used in the future. Some of the summaries retained jargon and results from this study will be used to optimize the prompt for an expanded study. In conclusion, a patient-friendly summary derived from a medical note using a large language model prompt was helpful to patients, and found to be useful for providers

**Author Summary:** As medical oncology providers, our new patient consultation appointments often require disclosing the diagnosis of a cancer, and a discussion on prognosis, complex treatment plans, the potential for significant side effects, and a number of tests/procedures that are required prior to initiation of the care plan. Patients often benefit from friends or family who take notes during an appointment, however this is not always possible. Technological advances in natural language processing with large language models such as Chat GPT allow for translation of medical language into plain language. In this study, we used a prompt to rewrite a medical note into a summary of the patient’s oncologic diagnosis and care plan. We then provided this summary to patients and provider to assess their feedback on the value of these summaries. We found that both providers and patients found these summaries to be accurate and understandable. Both groups recommended further development of these summaries. We intend to optimize our summary production for future studies using findings and feedback from this project.

## 1 Introduction

Large language models (LLMs) such as the generative pre-trained transformer (GPT) are powerful tools capable of natural language processing and bring great potential to improve upon the delivery of health care[1,2]. A strength of this technology is its ability to interpret the meaning of written sentences and to provide summaries of complex inputs. By using these tools to interpret medical notes, it is possible to create a written summary without medical jargon providing a patient-specific summary[3].

Outpatient medical oncology consultations frequently include the discussion of life-threatening diagnoses, toxic medications with complex administration schedules and multiple options for formulating the optimal treatment approach. Evidence demonstrates that patient retention of information discussed in an initial consultation can be poor. Recall may be approved with support members present, or additional medical services provided by nursing, social work, nutrition, and other services[4-6]. General handouts such as chemotherapy information, or dietary recommendations can help, but are not patient specific. The medical note in the United States is available to most patients through the Open Notes initiative, however documentation often uses technical terms that make them inaccessible to most patients[7]. These notes contain a lot of patient specific information, such as treatment recommendations, additional testing to be done, as well as cancer stage and prognosis, which is of value to patients .

In this study, we sought to evaluate the capability for an LLM to provide an accurate summary from an oncology in a reliable manner. This was a pilot study designed to assess both patient and provider satisfaction with the output.

## Results

Between December 2024 and June 2025, twenty-seven patients enrolled on this study. Four patients were lost to follow up prior to a second appointment (Three for transfer of care, one deceased), and in all cases patients were taken off study so surveys have not been completed by patient or provider. An additional 8 patient surveys have not been completed either due to delays in the consultation noted being prepared prior to the follow-up; delays in providing a completed summary to the treating provider in advance of the appointment; or the follow-up being a telehealth/remote visit. When possible, attempts were made to provide the summary and survey at subsequent follow-up. A total of 23 provider surveys were completed. Fifteen patient surveys were completed. No summary was rejected for being inaccurate by the PI or treating physician. Two outputs had excessive technical terms and in these two cases, the LLM was prompted to re-write with less jargon.

The patient survey reported a mean 4.6 (out of 5) for accuracy, 4.8 for clarity and 4.4 for helpfulness of their summary, with replies of agree or strongly agree in 100%, 100%, and 87% on each topic respectively (Figure 1). 93% of the patients would recommend similar summaries to be used for other patients or by other specialties.. Patients reported satisfaction with the clarity and helpfulness of the tool. One patient noted concern of the staging being written as T4, which was misconstrued by the patient as stage IV.

**Figure 1.**
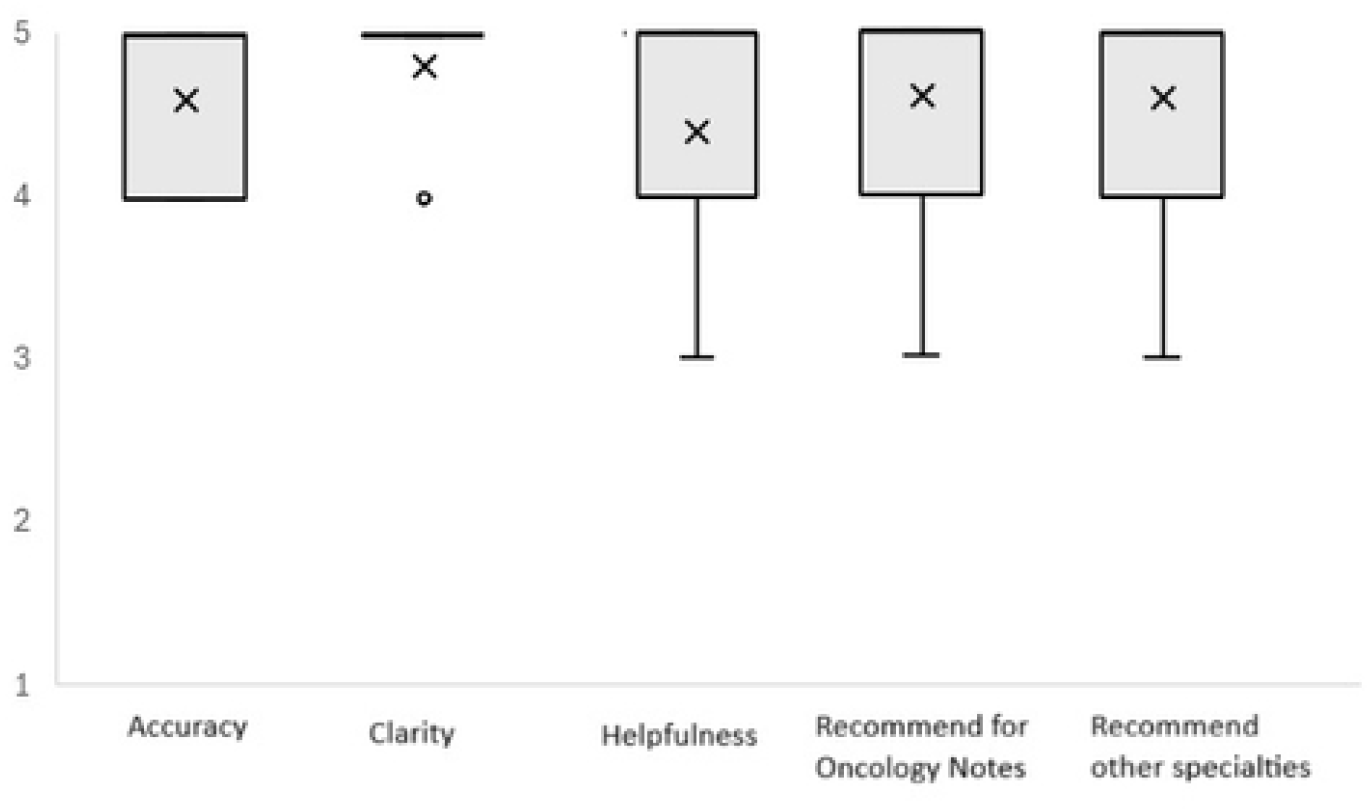
Box-and-whisker representation of patient survey results. “X” denotes mean score for each category of question, “o” denotes outliers, the grey box representing the middle 50% distribution, and lines representing 5%-95% range of responses.

The provider survey reported a mean 4.8 (out of 5) for accuracy, 4.4 for clarity, and 4.6 for empathy of the summary output with replies of agree or strongly agree in 98%, 91%, and 96% for each topic respectively (Figure 2). All providers (100%) reported that they would want to see similar surveys for oncology consultations in the future and 85% of reports thought such a summary would be helpful in other medical specialties. Some overly technical jargon was noted as an area for improvement. Initially the LLM was prompted to provide a ‘statement of support’ however after initial provider feedback, this was removed allowing the output to better reflect an individual provider’s voice and style. Sample comments from providers and patients are included in table 1.

**Table 1.**

| **Provider**<br>**Comments** |  |
| --- | --- |
|  | *Information is nicely summarized and succinct* |
|  | *Jargon... Could replace poorly-differentiated with different term* |
|  | *Good summary of side effects and secondary issues.* |
|  | *Hits the nail on the head - accurate and succinct* |
|  | *Easy for patients to understand. I liked the section headings* |
|  | *Organized and clearly presented the treatment plan* |
|  | *Included jargon such as 'sentinel lymph node biopsy', 'ovarian suppression'.* |
| **Patient**<br>**Comments** |  |
|  | *All aspects of this procedure were well laid out* |
|  | *The summary is very useful to refresh my memory where so much information has been discussed* |
|  | *I think every specialty should offer this* |
|  | *The doctor was clear in the oral communications but it is nice to have a summary that does not include a lot of medical terms* |
|  | *It gave the importantant information without the additional jargon and unnecessary details* |

**Figure 2.**
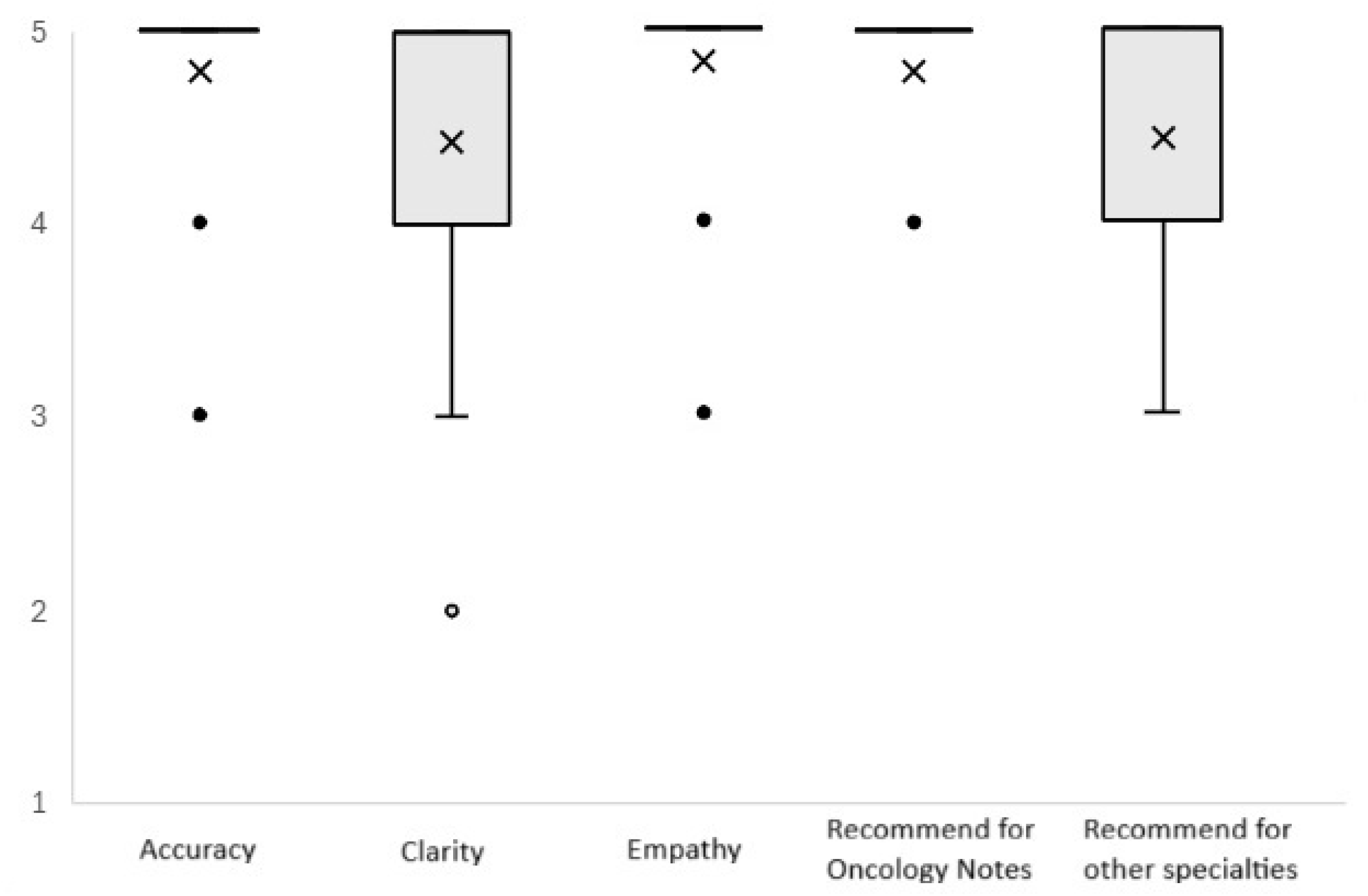
Box-and-whisker representation of provider survey results. “X” denotes mean score for each category of question, “o” denotes outliers, the grey box representing the middle 50% distribution, and lines representing 5%-95% range of responses.

As noted, two summaries were found to have retained a high number of technical terms. For both cases, the AI was asked to re-write the summary without technical jargon and the output was reviewed. For these two cases, the provider was able to review both summaries and the patient was provided with the second output. Both events arose after the LLM upgrade from GPT 3.5 to 4o. Interestingly many of the terms in these two outputs (peritoneum, neuropathy, shorthand terms such as “CT” (Computed tomography) or “5-FU” (fluorouracil)) were simplified into more straightforward terms in preceding summaries on a first pass.

There were some repeat errors noted in the summaries. Cancer stage was sometimes reported only partially, such as the local extent of spread (T stage), rather than the summary stage which considers the T stage along with nodal or distant involvement. This appeared to depend on how individual providers reported information in their note. A few frequently used terms, notably sentinel lymph node biopsy and axilla, also were seen on repeat reports, all of which were more frequently encountered after the upgrade to GPT-4o.

## Discussion

This was a pilot study to assess the reliability of a LLM oncology patient-friendly summary as well as to obtain provider and patient feedback. Overall, these summaries were accurate to the documented note and we did not observe any hallucinations or inaccurate information beyond what was requested of the AI. Despite technical terms being present in some of the summaries, this did not seem to greatly impact patient reviews and is likely to be addressed with modifications to the primary prompt prior to a further expanded roll out of the generated summaries.

There are many potential benefits of these studies. By having a written summary of a discussion, this may help reduce the number of follow up calls for clarification or reduce missed appointments for scheduled imaging studies or infusions. They could increase patient trust in providers by making documented information more accessible to patients. We would anticipate that patients presenting to consultations alone to benefit from a written summary. For patients with support members taking notes, a second written summary may help reduce risk of inaccurately recorded information. Future studies are planned to objectively evaluate the benefits to the health system and patient experience. The Open Notes initiative based on the 21st Century Cures act has advocated transparency between medical records and patients[7]. Medical jargon obscures this transparency and AI-driven summaries provide an opportunity to improve further upon this ideal.

There are other initiatives to extract medical data and documentation into a provider friendly summaries[8-10]. One major area of focus is to distill an entire hospital stay into a AI-derived discharge summary. Such a summary is complicated by the volume of data that needs to be analysed and prioritized[11]. The summary from an oncology note requires much less nuanced prompting as the data in a consultation note is much briefer than the innumerable documents that come from a multi-day hospital admission. This prevents hallucinations and is much less likely to miss any critical information.

Although the initial summaries were well received by patients and providers, there are a few areas that can be optimized, such as with differentiating between TNM staging and summary stage[12]. In addition, the LLM was prompted to summarize anticipated toxicities from chemotherapy regimen recommended. As providers document details of stage or expected toxicities in different ways, this does lead to inconsistencies in output. Although allowing for some variation in output to match a provider documentation style and voice can be a strength in this summary, certain things such as expected side effects from a specific regimen can likely be standardized. We intend to use the capabilities of LLMs to pull from external references on chemotherapy toxicities and to calculate summary stage when not explicitly stated, using the available data.

One noted limitation was related to the emergence of increasing technical terms. When this study was designed, Doximity was the only HIPAA compliant LLM that was available at our institution. This is a tool used predominantly by physicians and predominantly trained on medical journals and the nature of LLMs are to learn from prior prompts and outcomes[13]. It is possible that the predominantly technical use of the tool by other physicians may have biased the study at the time of update, although this cannot be confirmed. We will be further optimizing our prompt and exploring the burgeoning options for HIPAA compliant LLMs within our institution to address this issue in the future.

Our study has several limitations. Given the small number of patients studied, there is a possibility that observer bias may have impacted provider documentation style, as well as patient reviews. LLMs are upgrading at a rapid pace, and continued quality review will be needed to ensure future upgrades do not lead to poorer quality summaries. The current study used an external LLM tool, which limits the ability to automate the process, however these tools are likely to be increasingly integrated into health systems in the coming months and years. We will also develop an improved process to get the summary to a patient in a convenient manner, potentially directly through the EMR, to reduce the loss to follow up and delay to summary delivery (prior to subsequent visit rather than during the visit).

## Conclusion

In summary, LLMs are capable of creating a clear, accurate summary from a medical oncology consultation note. In this pilot study, Patients and providers both found these summaries helpful and capable of improving patient understanding of complex care plans. Future efforts should seek to identify the benefits of these summaries to patient care and health system quality metrics.

## 2 Methods

### 2.1 Setting and Patients

This study was conducted at a university-based medical oncology practice in Worcester, Massachusetts. Patients included those who present for an initial consultation to providers with subspecialty expertise in genitourinary, gastrointestinal, hepatobiliary, breast, or thoracic oncology. At the conclusion of the visit, this study would be offered to patients. As the purpose of this study was to assess the ability to summarize treatment plans, providers were encouraged to recruit patients who had some form of systemic therapy recommended. All patients provided written consent to be enrolled onto study

### 2.2 Procedures

After enrolling in the study, the completed medical note was obtained by the principal investigator. Identifying information was censored from the document. The remainder of the medical note was input into a Health Insurance Portability and Accountability Act (HIPAA) compliant LLM (Doximity, powered by ChatGPT). During the study, Doximity upgraded its GPT-3.5 to GPT-4o. This LLM was selected as it was the only readily available HIPAA-compliant tool with institutional approval. The LLM was prompted to summarize and include the diagnosis, stage of cancer, treatment plan, any symptom management recommendations, and further diagnostic tests. These topics were selected as considered standard aspects of most visits. The LLM was also prompted to present the information in plain language and to limit harsh terms. The prompt was engineered using a series of sample oncology consultation notes from fictitious to optimize output prior to initiation of the study[3].

Once the summary was produced, the principal investigator and the treating oncologist would review the output to ensure no factual errors were present. The treating oncologist was instructed to complete a survey using a 5-point Likert scale (Surveys available in Supplement) to rate the output’s accuracy, presence of jargon, degree of empathy, and overall satisfaction. At the patient’s next follow up; while awaiting the provider, they received the summary and a similar survey for completion. Patients were allowed to keep the summary if they wished.

### 2.3 Ethical Considerations

This study’s protocol was approved by the University of Massachusetts Chan School of Medicine Investigational Review Board. This study was conducted in accordance with the Declaration of Helsinki, with all patients provided voluntary written consent and were not reimbursed with financial or material compensation. All collected data, prior to submission into the LLM, were anonymized with personal identifying information censored.

### 2.4 Statistical Analysis

A descriptive analysis of aggregate Likert survey data is provided and with mean calculated with Microsoft Excel. Scores were reported on a scale of 1 (“strongly disagree”) to 5 (“Strongly agree”). Bar whisker plots are displayed for visual representation.

## Data Availability

All data will be made available in supporting data without restriction with publication in supporting data file

## Acknowledgements

The authors would like to acknowledge the assistance of Allen Cheng who provided valuable insights on large language model selection and prompt development. We also are grateful for the generous assistance by Preya Patel to ensure patient enrollment met regulatory requirements.

